# Tracking the Dynamics and Allocating Tests for COVID-19 in Real-Time: an Acceleration Index with an Application to French Age Groups and Départements^*^

**DOI:** 10.1101/2020.11.05.20226597

**Authors:** Christelle Baunez, Mickael Degoulet, Stéphane Luchini, Patrick A. Pintus, Miriam Teschl

**Affiliations:** Aix-Marseille Univ, CNRS, Institut Neurosciences Timone.; Aix-Marseille Univ, CNRS, Aix-Marseille School of Economics.; Aix-Marseille Univ, CNRS, EHESS, Aix-Marseille School of Economics.

**Keywords:** COVID-19, Indicator of Epidemic Dynamics, Acceleration Index, Real-time Analysis, Sub-National Allocation of Tests, France

## Abstract

An acceleration index is proposed as a novel indicator to track the dynamics of the COVID-19 in real-time. Using French data on cases and tests for the period following the first lock-down - from May 13, 2020, onwards - our acceleration index shows that the ongoing pandemic resurgence can be dated to begin around July 7. It uncovers that the pandemic acceleration has been stronger than national average for the [59 − 68] and especially the 69 and older age groups since early September, the latter being associated with the strongest acceleration index, as of October 25. In contrast, acceleration among the [19 − 28] age group is the lowest and is about half that of the [69 − 78], as of October 25. In addition, we propose an algorithm to allocate tests among French départements, based on both the acceleration index and the feedback effect of testing. Our acceleration-based allocation differs from the actual distribution over French territories, which is population-based. We argue that both our acceleration index and our allocation algorithm are useful tools to guide public health policies as France enters a second lock-down period with indeterminate duration.

**JEL Classification Numbers:** I18; H12

## 1 Introduction

The current COVID-19 pandemic not only caught most of the world by surprise, it also highlighted the crucial aspect of uncertainty under which all societies live since the first mentioning of the new viral infection at the end of 2019. This uncertainty however not only concerns the novel SARS-Cov-2 pathogen and how it spreads, its effects on human health, the best possible treatments, when a vaccine will be available and how efficient it will be, to just cite some of the concerns. There are at least two more uncertainties which receive less attention. One is parameter-uncertainty of typical SIR (Susceptible-Infected-Removed)-models that are widely used to predict the evolution and consequences of the pandemic, and the other is data-uncertainty.^1^ While a continuous effort is put into remedying and improving data collection, because this is, after all, the first and foremost important source of empirical knowledge on which all else is built, the parameter-uncertainty is intrinsic to modelling strategies and thus much less prone to any easy solutions.^2^ A key point of this paper therefore is to say that given these uncertainties, what is of crucial importance to know, here and now, for public health specialists and decision makers and other people alike, is whether harm is accelerating or whether measures that are put in place to control the pandemic are contributing to curbing the spread and thus to decelerate harm (see Taleb [11]). In the case of a pandemic, harm can be defined, for example, as the number of confirmed cases. However, simply plotting the number of positive cases over time, as it has been done on many governmental sites, is not the right way of understanding the dynamics of harm. For example, the knowledge of positive cases depends on testing and hence cases will very much depend on the underlying testing strategy put in place. But this is exactly our point: to say something meaningful, we need to put cases and tests in relation to each other. That is, to understand whether harm, defined as the number of cases, is accelerating or decelerating, we need to plot cases against tests and not to consider them separately.

Although this might be seen as just another tool to visualize the data, we show that such a way to organize the data is extremely useful to uncover an index of acceleration, which turns out to be a simple and plain scale elasticity at the end date of the sample. It tells us how much additional cases are detected given additional tests, in percentage of the total of both variables attained at the date for which one has the last vintage of data. We argue that updating such an acceleration index in real time provides very valuable information and is an essential tool to apprehend the pandemic uncertainties with which we currently live. It is also dealing with parameter-uncertainties and this simply because we do not use any. Instead of engaging in assumptions and probability attributions to ex-ante uncertain events, we use the data that is currently available, which is the best we have, to shed light onto the dynamics of the pandemic and the harm it produces. Our aim in this paper, to be clear, is not to provide a detailed analysis of the pandemic dynamics but to clarify what can be done with available data. Our paper is of conceptual nature, proposing a new way of apprehending and looking at uncertain events (Degoulet et al. [4]). We present a real-time operational tool to complement epidemiological SIR models, which are limited in their insights from an ex-ante perspective.

In what follows, we present in more details our novel indicator to track the pandemic: the acceleration index (section 2.1). Based on this indicator, we show that after a period of deceleration since the end of the first lock-down in France in mid-May, using the data provided by the French Public Health Agency (Agence Santé Publique France), we can observe acceleration emerging from about July 7 onwards, which is about two weeks earlier than other indicators, for example the positivity rate, which was introduced by the French Public Health Agency over the summer to assess the available data.^3^ In a period where time counts and early indications of acceleration should be used as warnings to rapidly prepare health care facilities and to reconsider public health measures in view of increasing the capacities to trace and thus to limit the transmission of the virus, this is essential information and thus an important advantage of our indicator.

Another advantage of our new indicator is that it can be used to zoom in on particular groups or geographical areas (section 2.2). Indeed, the indicator can be more helpful the more fine-grained the level at which it is applied. We present a first analysis of the pandemic dynamics for different age-groups and regions in France. Regarding age groups, what we find is that the acceleration over the last two months has been driven more strongly by the age group of people older than 59 than by the age groups between 10 and 28. In particular, the elderly aged 79 or more have not been preserved from this new acceleration. This is important information as it seems that it is not professional or school and educational activities that drive the pandemic’s acceleration. It raises the important and serious question of why harm accelerates exactly at the age groups which are most at risk. On the other hand, it shows that one of the hot-spots for virus transmission at the beginning of the pandemic, nursing homes, has not quite adapted to the new living constraints.

As for the spatial distribution of the dynamics, we present maps with French “départements” which indicate the evolution of the differences in acceleration across France from about the period of the end of the first lock-down in mid-May, to the beginning of the second lock-down, end of October. We also look more closely at the acceleration index for the four largest “départements” (Bouches-du-Rhône with Marseille, the North with Lille, Paris and Rhône with Lyon) and compare in particular the acceleration within these areas of the two age groups which are facing a greater acceleration over the last two months than the overall acceleration in France.

Finally, based on our indicator, we propose an algorithm for an endogenous testing strategy that takes into account the differences in acceleration across space and age groups (section 3). Any test strategy combines two aspects: information about the virus circulation and the expected feedback that testing implies, in terms of isolation and contact tracing most importantly. We propose a parsimonious algorithm to allocate tests across age groups and space, based on both our acceleration index and the average positivity rate, and on the extent to which tests reduce the virus propagation. Because the latter feature can never be measured with certainty, we argue that any allocation of tests should also reflect the beliefs of (benevolent) public health authorities and experts and we add a parameter to the analysis to take into account this dimension explicitly. We show that our acceleration-based allocation of tests differs from the actual distribution of tests, which has essentially been determined by population size. But our analysis shows that population size is not necessarily the criterion that affects acceleration, at least not all the time. Hence, to the extent that testing is accompanied by contact tracing and efficient isolation, this observation means that allocating tests according to where the virus accelerates the most would be a better way to control the pandemic.

## 2 Tracking the COVID-19 Dynamics in Real-Time

### 2.1 Daily and Average Positivity Rates, and the Acceleration Index

Accurate and reliable information about how pathogens spread over space and time is of first-order importance to fight epidemics and to properly design efficient public health policies. What is striking in the case of COVID-19 is that, at least in North American and European countries, the main indicators to track the *dynamics* of the pandemic have been rather coarse and are still limited. To go back to the early stages of the pandemic outside China and neighboring countries, from March to the summer of 2020, the main data used by public health authorities and available to the general public to track the pandemic has been summarized by the time series of the number of positive cases over time.

In Figure 1 are shown, in panels (*a*) and (*b*) (top row), such numbers for France, starting May 13 which is when the lock-down for the entire country ended.^4^ Panel (*a*) shows the daily number of people who have been tested positive to COVID-19 for each day, while panel (*b*) shows the daily but cumulated number of such people over time. It is worth noting that this type of graphs was all there was to visualize the dynamics of the pandemic, at least until late in Spring. In the case of France, the agency in charge of producing public data related to COVID-19, Santé Publique France, initially communicated in terms of number of cases over time only.^5^

**Figure 1:**
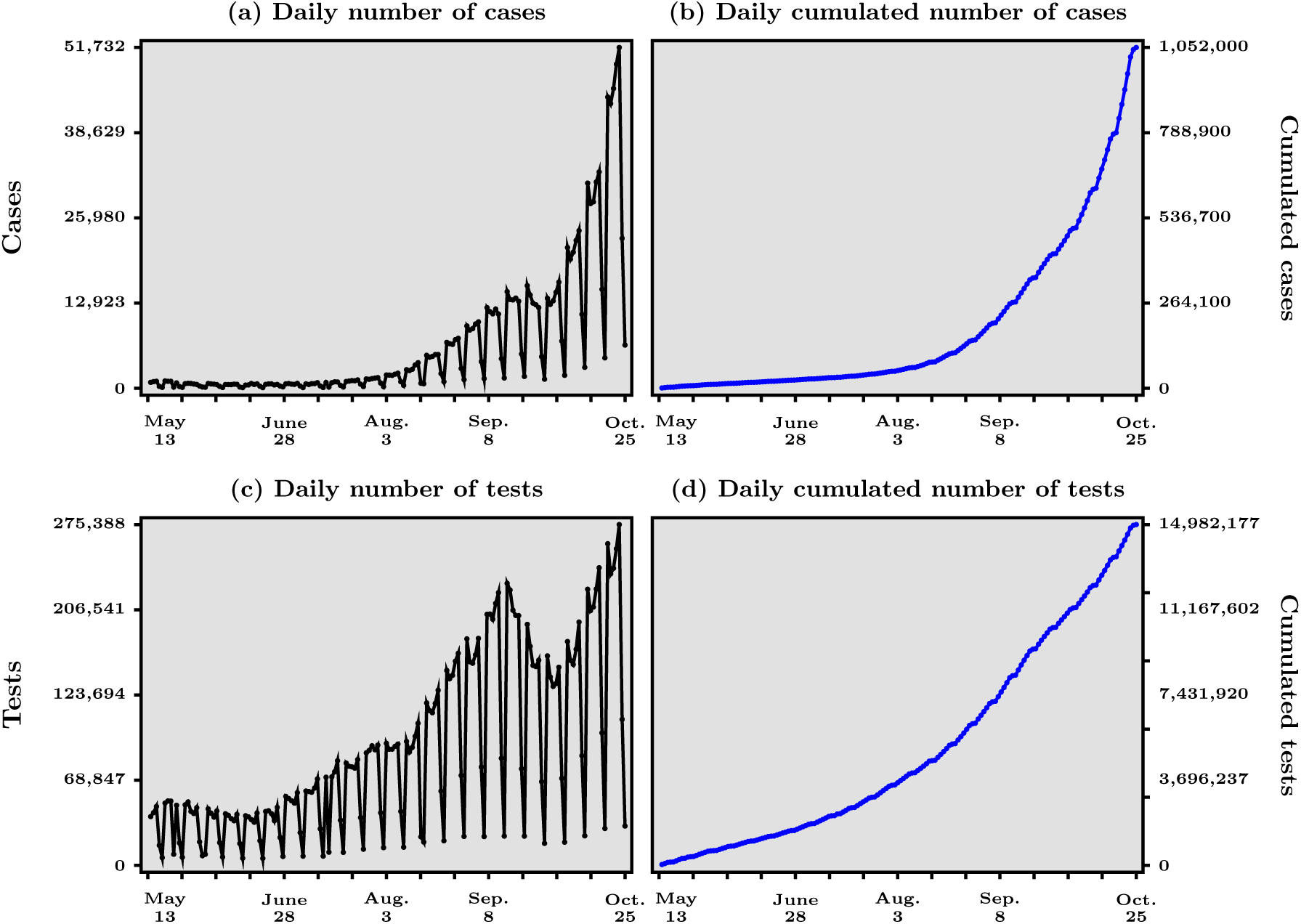
Numbers of confirmed cases and tests, daily and cumulated, for COVID-19 in France, May 13 - October 25, 2020. Data source: Agence Santé Publique France

The problem with the information contained in the graphs of the first row in Figure 1 is that they would be a fine summary of the pandemic’s dynamics if the number of tests performed over a unit of time was roughly constant. In that case, the evolution of the number of new cases, given a constant over time number of new tests, would capture well *how fast* the virus spreads. However, as shown in panels (*c*) and (*d*) in Figure 1 (bottom row), this assumption is far from correct in practice, and this is not only true for France because of several reasons that has concerned most countries.^6^ In the period after the end of the generalized lock-down in France, when the WHO message to “test, test, test”^7^ was becoming effectively received in France, one sees from panel (*c*) in Figure 1 that the number of tests performed each day is far from constant and has in fact been trending up most of the time, not surprisingly given the level of unpreparedness of the French government at the beginning of the pandemic.

It is obvious that the dynamics of the number of cases is strongly related to the dynamics of performed tests. After all, diagnostics are the only, albeit imperfect, way to confirm that people are positive to COVID-19.^8^ However, observing each independently (side by side) may give only a blurring view at best, and even a confusing sense, of the extent to which the pandemic accelerates. A manifestation of how confusing that coarse piece of information can be, if not properly organized, is the statements often voiced that one should not be worried to observe more positive cases as long as more tests are performed as time passes. After all, more tests imply more cases, almost by definition and this basic fact should not be taken at face value to indicate that the pandemic is worsening.

We argue that such reasoning is wrong and that the correct understanding, in terms of measuring the *acceleration/deceleration* of the pandemic, is gained if a scatter-plot of the number of positive cases against the number of the tests is used in real time, instead of the panels in Figure 1. This is the first contribution of this paper, in our view, and it is presented in Figure 2. Before presenting our preferred way to organize the data, a few remarks are in order.

**Figure 2:**
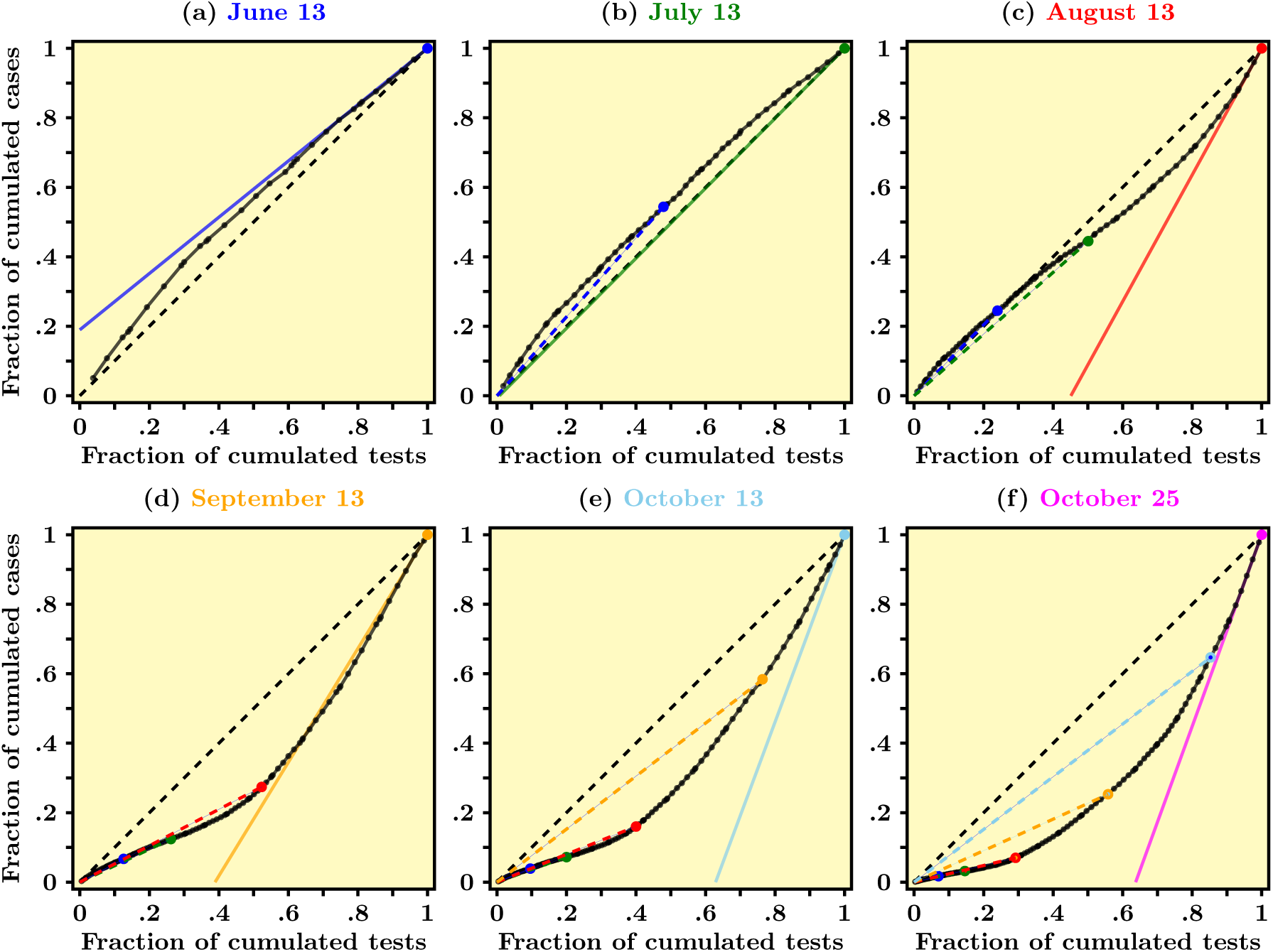
Scatter-plot of cumulated daily cases against cumulated daily tests for COVID-19 in France at different dates - Data source: Agence Santé Publique France. Data normalized using min-max re-scaling. The dashed black line represents the diagonal. The slope of colored solid lines in each panel is a visualization of our acceleration index. The dashed colored lines represent the chords associated with previous dates.

First, we use the cumulated numbers of daily cases and of daily tests as a way to keep track of the size and overall prevalence of the pandemic. Second, our proposed scatter-plot has merit only if it is *updated in real time*, say, at daily frequency, so as to add information about additional tests and cases. This immediately raises the question of how data should be normalized, given that the number of data points obviously increases with time. We choose a simple and rather innocuous re-scaling method known as min-max normalization,^9^ which in our case amounts to divide all historical values by the last data point figure. As a result, our (all positive) normalized data values are contained within the unit interval (0, 1) so that our scatter-plot lies in the unit square.

In Figure 2 we present our proposed scatter-plot, which is in our view a very useful way to visualize the dynamics over time of the number of cases *in relation to the number of tests*. As alluded to above, there is a causality arrow going from the latter to the former, so we plot cases against tests, all in cumulated values. For example, panel (*a*) of Figure 2 graphs all data points from March 13 to June 13, 2020, each point/dot representing a particular date. On the *x*-axis, all numbers are between zero and one due to min-max normalization and a particular value gives the fraction of tests performed that day, say, *t*, out of the total cumulated number of tests performed up to end date *T*, say. For example, 0.8 means 80% of the total amount of tests cumulated from date 0 (May 13, 2020, in our case) to date *T* (June 13, 2020). Similarly, along the *y*-axis we report the cumulated number of cases for each day *t* ≤ *T*, as a fraction of the total number of cases at date *T*. All such normalized data points fill in a solid black curve which then represents the actual data over time. Starting at the origin and moving along the solid curve in the north-east direction means moving forward through time, since we use values that add up over time.

The dashed black line in panel (*a*) of Figure 2, on the other hand, is the first diagonal and it turns out to be an interesting benchmark if one is interested in capturing the acceleration/deceleration of the pandemic. Counter-factually, If the dashed diagonal would represent the real data, that would tell us that the virus spreads according to a linear pattern whereby, say, 20% of the total amount of tests accounts for 20% of the total number of positive cases, and so on. And this coincidence would hold at all dates between zero and *T*. In other words, the pandemic neither accelerates nor decelerates since a given proportion of tests account for the same proportion of detected cases for all the data period, that is, over time.

Panel (*a*) of Figure 2 shows that this has in reality not been the case for the period May 13-June 13, 2020, or for all the period covered by our analysis for that matter. More precisely, the solid line, which represents actual data, turns out to be entirely lying above the dashed diagonal, which should be interpreted in the following manner. First, the blue line representing the tangent to the black curve at the end point (1, 1) turns out to have a slope smaller than one (this is easy to see since the slope of the dashed diagonal equals one by definition). This means that between the day right before *T* and day *T*, the observed fraction of *new* tests (out of the total number of tests to this day) has been associated to a *smaller* fraction of *new* cases (out of the total of cases to this day). We then want to say that at date *T* the pandemic is decelerating. Now going backward from date *T* into the past (that is, moving from the farthest north-east corner toward the origin along the solid curve), one sees the “slope” of the tangent to the solid curve going up as time moves backward. In other words, the slope of the tangent to the solid curve has been decreasing from date 0 to date *T*, which is an equivalent way of saying that the solid curve is concave: for a given fraction of tests, lower and lower fractions of cases have been detected over time. This indeed captures in a visual fashion the fact that the pandemic has been continuously decelerating between May 13 and June 13. This observation indicates that France has somehow benefited from the lock-down period in the sense that the circulation of the virus, in the aggregate, slowed down.

Unfortunately, this state of affairs was not to persist. Panel (*b*) in Figure 2 shows, using color coding, that a month later, on July 13, the slope of the solid curve at end date (the green line) was about one - the green line almost coincides with the dashed diagonal. In other words, the pattern of the pandemic over time switched from decelerating to roughly linear, between mid-June and mid-July. Quite importantly, comparing panel (*a*) to panel (*b*) suggests that the slope at end date had started to increase and has crossed unity between June 13 and July 13. While comparing panels (*a*) and (*b*) suggests the end of deceleration, panel (*c*), dated August 13, clearly shows that *acceleration* was then taking place: the slope at end date is about two. In addition, the scatter-plot at later dates reveal that acceleration is still underway, to this day (October 25, 2020).

One important conclusion is that Figure 2 delivers an indicator in real time of the pandemic’s dynamics, in the sense that it helps to visualize whether the pandemic accelerates or decelerates at any date. The property that the pandemic accelerates when the slope of the solid curve at end date is larger than one, and that it decelerates when the slope is smaller than one, is in fact no surprise. As shown in Appendix A, this slope is essentially an *elasticity*: its value gives the variation of cases detected by a given variation of tests, in proportion to levels of (cumulated) cases and tests attained at end date.^10^ For example, a value about two as in panel (*c*) of Figure 2 means that a given proportional increase in the amount of tests accounts for a proportional increase in the number of cases that is twice as much. One can therefore think of this elasticity as an *acceleration index*: acceleration happens when the elasticity is larger than one, otherwise the pandemic decelerates. In the razor’s edge case when the elasticity is exactly one, a linear pattern emerges so that the pandemic neither accelerates nor decelerates.

In each panel (*b*) − (*e*) of Figure 2, we also report the points corresponding to the previous dates associated with the other panels. For example, the blue dot in panel (*b*) corresponds to the situation at June 13. Similarly, the red dot in panel (*d*) corresponds to August 13. Interestingly, our scatter-plot helps also visualizing the pandemic’s dynamics in a retrospective fashion, to the extent that one can visually track not only the slope of the derivative to the solid line, but also the slope of the cord that links the origin to the point corresponding to the date under scrutiny and we now turn to this subtle point.

More specifically, although Figure 2 and the depiction of our acceleration index are very helpful to visualize in real-time whether the pandemic accelerates at end date, those two visualization tools have their own limitations and must be refined if one is to do a retrospective analysis of the pandemic using the information in the scatter-plot and not only the elasticity at end point.^11^ The way we see it is that the scatter-plot should be updated in real time to visualize where the pandemic goes in terms of acceleration/deceleration. We can in particular infer each day, from the most recent data point, the slope of the tangent to the solid curve, the value of which is what we called the acceleration as explained above. But such an index by definition combines two types of information: variations (between two successive dates in our example) and levels at final date. To use the notation of Appendix A and if we denote the elasticity/acceleration index at end date *ε*_T_, the following decomposition holds:

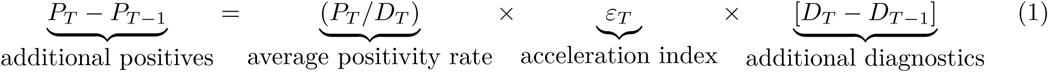

where *P*_*T*_ and *D*_*T*_ denote respectively the cumulated numbers of positives/cases and diagnosis/tests at date *T*, and similarly for date *T* − 1. If we define from equation (1) the daily positivity rate to be the ratio between additional positives and additional tests, that is, what we have called earlier the tangent’s slope, then *the above decomposition relates the elasticity, the average positivity rate and the daily positivity rate*. More precisely, the elasticity is defined as the ratio of the daily positivity rate to the average positivity rate (much as the elasticity of a function is the ratio of its derivative to its average value). This fact suggests that looking at the average positivity rate alone is not informative enough, in the sense that this indicator does not say how much additional tests translate into additional cases, even though it is of course a fine measure of the average detection rate. It is a proxy for the prevalence of the virus in the population, a key parameter for herd immunity (see Britton et al. [3], Fontanet and Cauchemez [5]).

That said, there might be situations in which any pair of those indicators summarize all there is to know, the third indicator being somehow redundant. For instance, the case depicted in panel (*a*) in Figure 2 is simple in the sense that, at all dates, both the tangent’s slope and the average positivity rate go down over time, and that the former looks, on close inspection, smaller than the latter. From this property, one concludes that the elasticity is smaller than one. This is of course another way to express the property that the solid line is concave (hence above the dashed diagonal since the solid line goes through the origin and the upper-right edge of the unit square). In contrast, panel (*c*) in Figure 2 reveals that there has been a *change in curvature*, that is, a reversal from deceleration to acceleration, between June 13 and August 13. This can be seen from the fact that the least recent part of the solid curve is above the dashed line, while the most recent part is below it. In other words, the convex part indicates acceleration of the pandemic, while the concave part indicates deceleration at earlier dates. One difficulty that arises from such a change in curvature/motion is that the daily and average positivity rates may vary in different directions, with ambiguous implications about whether the elasticity stays smaller than one or, on the contrary, becomes persistently larger than one.

In panel (*c*) of Figure 2, one notices an inflexion point, when the solid curve intersects the dashed diagonal in its interior. At the date when this happens, the tangent’s slope/daily positivity rate starts to rise, whereas it was declining over time before that date. The average positivity rate, however, continues falling right after the inflexion point is attained, and it starts to rise only later. Eventually, both the daily and the average positivity rate do coincide, and when this happens the elasticity equals one. After that date, the elasticity stays larger than one. More specifically, imagine that the elasticity is smaller than unity and that the daily positivity starts increasing while the average positivity rate is still declining over time. It then becomes possible that the elasticity starts rising and eventually crosses the unit value and possibly stays persistently larger than one. The latter situation occurs whenever the daily positivity rate rises faster (or declines more slowly) than the average positivity rate.

Note that all this is of course reminiscent of what happens when the graph of a continuous and increasing function changes curvature, from concave to convex. We therefore conclude that while the colored line’s slope is an estimate of the elasticity at end point, looking back in time at past events from the perspective of date *T* through the lens of our scatter-plot requires to examine how both the daily and the average positivity rates vary over time, as shown in Figure 3.

**Figure 3:**
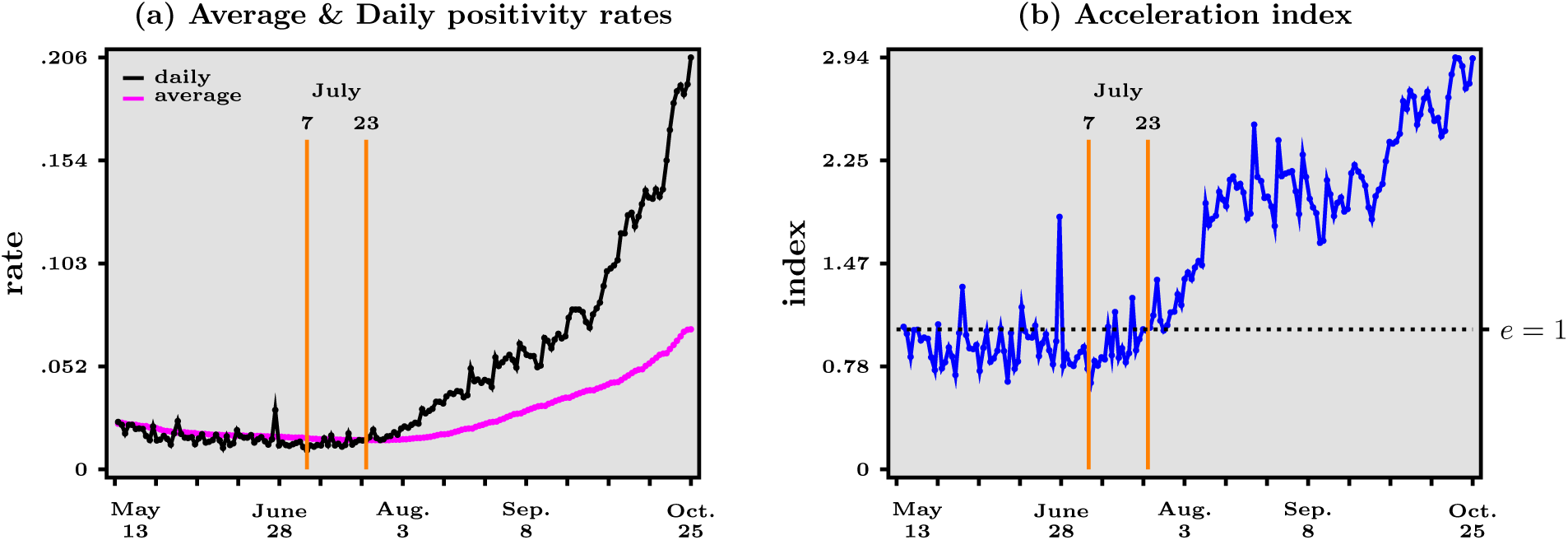
Main acceleration indicators - daily positivity rate (daily cases to daily tests ratio, black line) and average positivity rate (daily cumulated cases to daily cumulated tests ratio, purple line) over time (left panel), and acceleration index (ratio of daily positivity rate to average positivity rate, right panel, blue line) over time, May 13 - October 25, 2020. Dashed horizontal line in right panel represents when acceleration index equals 1. Data source: Agence Santé Publique France both the daily and the average positivity rates vary over time, as shown in Figure 3.

In Figure 3 we report over time the magnitudes of the three terms that can be inferred from the decomposition in equation (1). Remember that our objective is to detect whether the pandemic decelerates, accelerates or switches between those two types of motion. A striking feature in Figure 3 is that the acceleration index - in panel (*b*) - leads the average positivity rate - in panel (*a*) - in the sense that the former starts rising about two weeks before the latter starts rising as well. *In other words, the evolution of the acceleration over time appears to be an early warning of a future rise in the average positivity rate*. Eventually, in view of the reasoning above, one expects of course that the rising acceleration index crosses the unit value when the (rising) average positivity rate equals the (rising) daily positivity rate. The bottom line is that the decomposition that follows from the scatter-plot in Figure 2 may be useful to detect the early reversal from deceleration to acceleration. In the case of France, the elasticity starts rising about July 7, while the average positivity rate starts rising about two weeks later, on July 23, as indicated by the orange bars in Figure 3.^12^

To sum up, the important piece of information that one can derive from Figure 3 is that a reliable measure of the pandemic’s dynamics is the elasticity, which we can think of as an acceleration index. This elasticity leads the average positivity rate and is more dynamic than the daily positivity rates, which are other key statistics to follow. In practice, therefore, panels (*a*) and (*b*) should be used as real-time indicators of the pandemic’s dynamics. From the acceleration index and average positivity rate then follows the daily positivity rate, which is is still useful to translate the transmission dynamics back into levels (as opposed to normalized ratios).

From panel (*b*) in Figure 3, one infers that the period since the end of the first lock-down in France can roughly be split into four sub-periods. From May 13 to July 7, our acceleration index fluctuates around a value slightly lower than one, indicating that the pandemic is in a deceleration regime. Our indicator suggests that July 7 is the start of an ongoing acceleration regime in which the acceleration index increases, crosses the unit value on July 23, and continues to increase until about mid-August, when our acceleration index equals about 2. In that sense, this second sub-period can be seen as the fact that the pandemic worsens more and more over time. Then a new plateau occurs until around October 1st, after which the acceleration index rises again to reach a value of about 3 on October 25. Of course, the challenge that the country faces as the second lock-down period starts, on October 30, is to reverse that pattern to ensure that the acceleration index starts decreasing and eventually reaches values below one, which would indicate deceleration of the pandemic.

The simple case whereby both cases and tests grow exponentially, as detailed in Section B of the Appendix, is perhaps useful to interpret the four sub-periods just described. The first plateau associated with deceleration is compatible with a situation where cases have been growing at a slower rate than tests for a while. In that situation, the acceleration index is roughly constant over time, below unity. Then if the growth rate of positives becomes larger than the growth rate of tests, then the acceleration index starts rising and eventually crosses unity, to converge to a new and higher plateau associated with a value larger than one. Finally the most recent rise, starting early October, can be understood as a new jump of the growth rate of positives. Although the above interpretation stresses changes in the growth rate of positives, what matters more generally is how the ratio of growth rates - that of positives divided by that of tests - varies over time.

In addition, the scatter-plot also directly offers a measure of “global acceleration/convexity”: in panels (*d*)-(*e*) of Figure 2, the solid curve is located entirely below the dashed diagonal, which indicates that the persistent acceleration somehow “dominates” the earlier and short-lived deceleration.^13^

### 2.2 Estimating the Acceleration Index for Age Groups and Départements in France

Based on the analysis in the previous section, we propose to use the acceleration index, calculated from the end point of our scatter-plot, as a parsimonious way to track the pandemic’s dynamics. In this section, we show how useful such an index is to decompose the evolution of acceleration over age groups, first, and space, in a second step.

In Figure 4, we report the evolution over time of our acceleration index for nine age groups, all aggregated over space. In each panel, the blue line is a replica of panel (*b*) in Figure 3. For comparison against such an average benchmark, the orange line represents the acceleration index corresponding to the age group of each panel. One striking feature is that acceleration is underway for *all* age groups except people older than 79 years, as of October 25, 2020. In addition, the acceleration for each age group tracks rather closely that of the average for some groups but much less so or not all for others.

**Figure 4:**
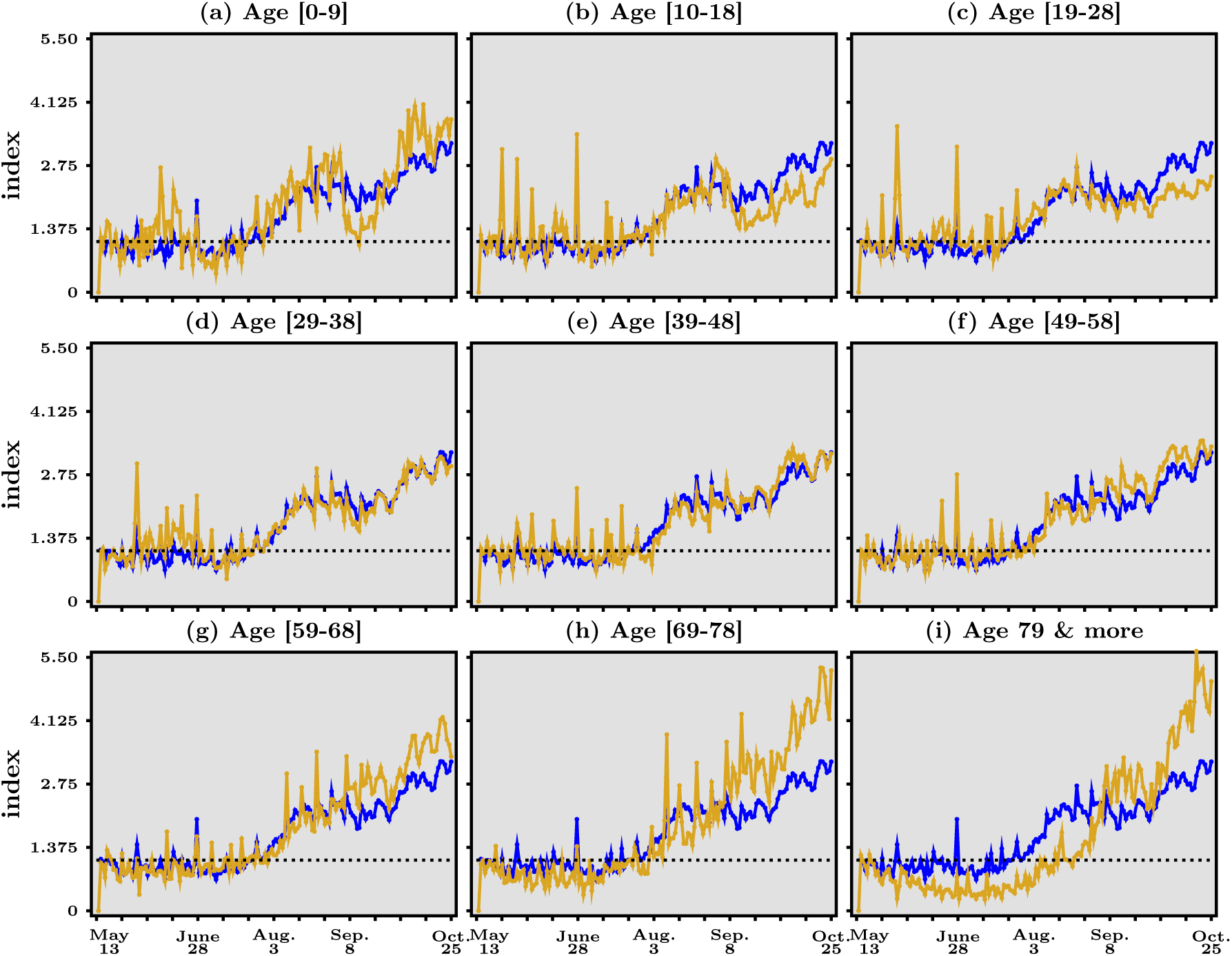
Acceleration index for different age groups, May 13 - October 25, 2020. In each panel, the blue line reproduces panel (*b*) in Figure 3, the dashed horizontal line represents when acceleration index equals 1, while the orange line corresponds to the age group specific to that panel. Data source: Agence Santé Publique France

To start with the former groups, the middle row in Figure 4 shows that people aged 29 to 58 experience acceleration in much the same way as the average over all age groups. This is not the case for younger and older groups. Children aged 10 to 18 and, quite spectacularly, young adults aged 19 to 28 seem to face a noticeable but slower acceleration since summer than the general acceleration. The latter group, in particular, is associated with an acceleration index that is maintained at intermediate levels around 2 in the last quarter of the data sample.

The fact that the share of confirmed cases is the second highest (22.9%) for the [20-29] age group (see Table 1 in Appendix C for incidence rates among age groups) may contribute to the view that youngsters such as college students have contributed a lot to the resurgence of the virus since last summer. This view does not agree with the data in Figure 4. The age group [0 − 9] has experienced ups and downs since last summer. In particular, the acceleration index has started to rise again quickly in early September across France, after it went down all the way to unity at the end of summer. The question here of course is whether the rapid increase has to do with the beginning of the academic year and the fact that young pupils did not wear face masks at school. However, another reason may be that younger kids have again been in more contact with their grand-parents, who take care of them some of the time while parents are working since the beginning of the school year. Yet in Figure 4, it is striking that the acceleration index for the age group [69 − 78] has risen importantly since the beginning of August, to attain the highest level at the end of the data sample, around 4! A similar, though slightly less strong rise, can be seen for people aged [59 − 68]. Hence maybe it is not the young kids who infect their grand-parents, but it is the other way around? Studies suggest that the “engines of the SARS-COV-2 spread” are household transmissions, which may support this hypothesis (see Lee et al. [9])).

**Table 1:**
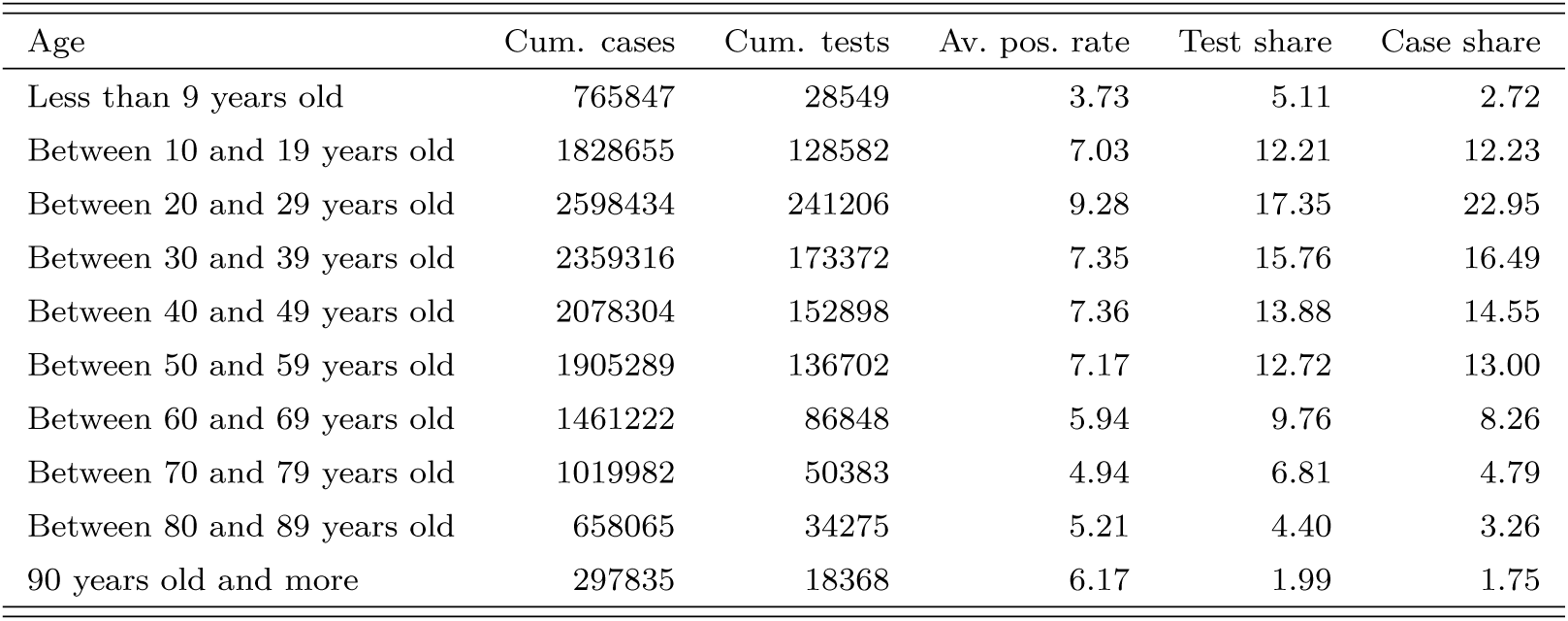
Some statistics for all age groups, as of October 25, 2020.

| Age | Cum. cases | Cum. tests | Av. pos. rate | Test share | Case share |
| --- | --- | --- | --- | --- | --- |
| Less than 9 years old | 765847 | 28549 | 3.73 | 5.11 | 2.72 |
| Between 10 and 19 years old | 1828655 | 128582 | 7.03 | 12.21 | 12.23 |
| Between 20 and 29 years old | 2598434 | 241206 | 9.28 | 17.35 | 22.95 |
| Between 30 and 39 years old | 2359316 | 173372 | 7.35 | 15.76 | 16.49 |
| Between 40 and 49 years old | 2078304 | 152898 | 7.36 | 13.88 | 14.55 |
| Between 50 and 59 years old | 1905289 | 136702 | 7.17 | 12.72 | 13.00 |
| Between 60 and 69 years old | 1461222 | 86848 | 5.94 | 9.76 | 8.26 |
| Between 70 and 79 years old | 1019982 | 50383 | 4.94 | 6.81 | 4.79 |
| Between 80 and 89 years old | 658065 | 34275 | 5.21 | 4.40 | 3.26 |
| 90 years old and more | 297835 | 18368 | 6.17 | 1.99 | 1.75 |

Equally remarkable is the group of people older than 79, which has experienced a catching-up in acceleration since mid-August, similar to - though later than - the dynamics of all other age groups. This suggests that the continued presence of COVID-19 does not seem to be under control in specialized nursing homes (the “EPHAD”). Given the large number of deaths among the very elderly at the beginning of the pandemic, this fact raises some serious concern.

Our analysis can also be applied to shed light on the local evolution of the pandemic. In Figure 5, we report in a map of France the acceleration index at the level of administrative division which is intermediate between the city and the region, called “département”. Red entries represent places where the pandemic accelerates (the elasticity is larger than one), while green entries represent decelerating départements. Our map does not necessarily coincide with the map very recently introduced by the European Centre for Disease Prevention and Control (ECDC) for Europe.^14^ In that map, green-orange and red lights depend on the incidence rate and the test positivity rate of a country but their combination of factors is not dynamic and does not have any clear conceptual foundation such as our acceleration-based index has. Their demarcation criteria between the different colours are set at a certain level, but it is not clear whether that level is sound from the perspective of our acceleration index.

**Figure 5:**
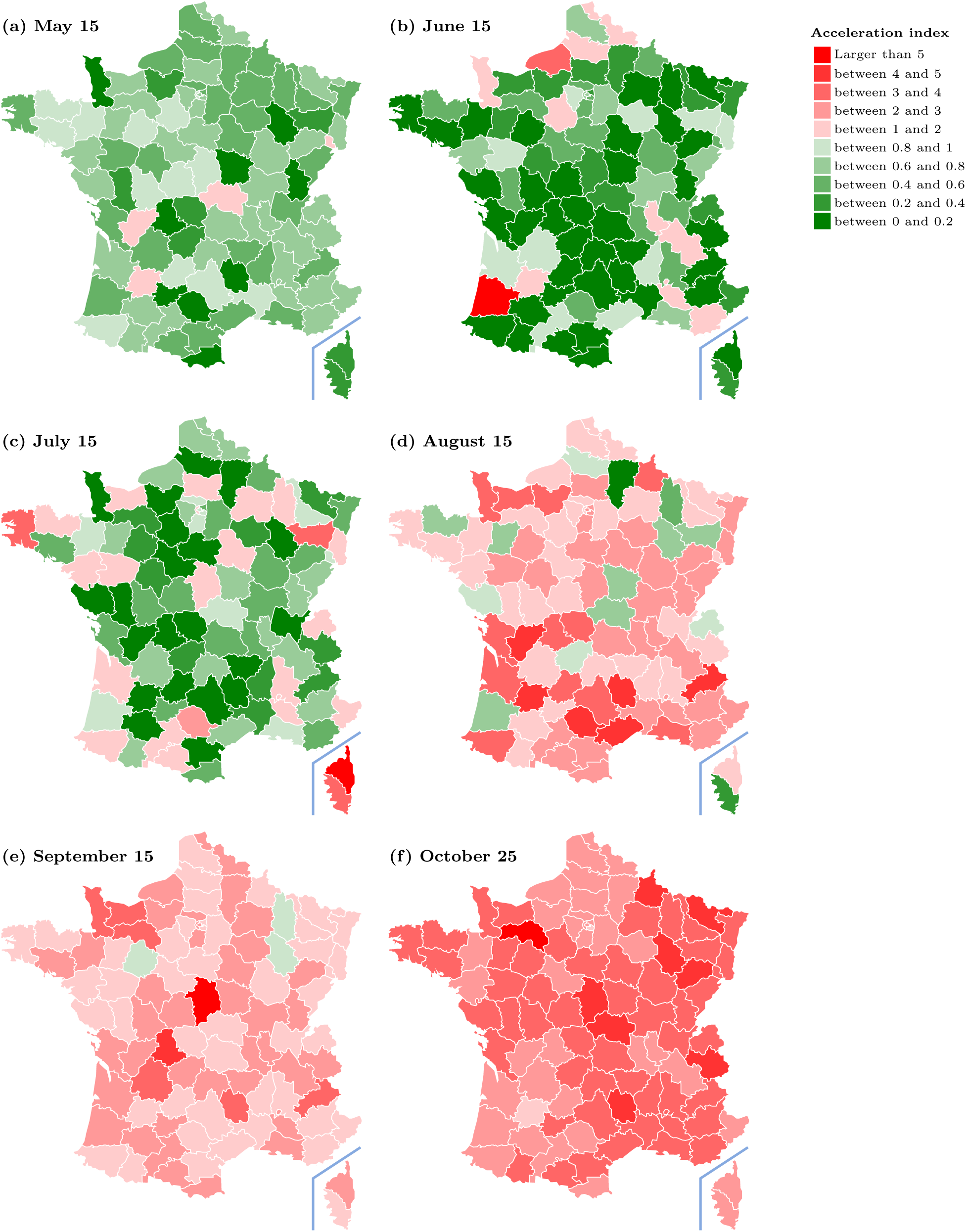
Acceleration index for French départements at different dates. Data source: Agence Santé Publique France

The upper left panel in Figure 5 depicts the level of the acceleration index for each département on May 15, that is, on the third day after the end of the lock-down period in France. While at that date, the pandemic is decelerating over almost all the country, the situation starts to gradually but dramatically change within the next two months and to reverse course over the summer break.

Between July 15 and August 15 (panels in second row), the country switch to a situation such that acceleration takes over the country. As of October 25, most département face an acceleration index larger than two, with no sign of reversal.

At the time when France enters a second lock-down period, lessons should be drawn about what went wrong in the period following the end of the first lock-down and we argue that it is important to analyse closely the reasons for local and group-specific increased accelerations. Knowing about local and group specific accelerations can be an important guide in investigating further possible transmission channels and networks, which would then allow to intervene specifically at a network and structural level if needed.

To give an example of such a fine-grained analysis, we dig a little deeper into the heterogeneity that is visible in Figure 5, and the accelerations we spotted in the different age groups. We report in Figure 6 our acceleration index for the four largest départements in terms of population, namely Bouches du Rhône, Rhône, Paris and Nord, which have been allocated more tests than smaller départements as the next section will show. Figure 6 features in panel (*a*) the dynamics for all age groups combined, while panels (*b*) and (*c*) depict that of kids aged [0 − 9] and of people aged [59 − 79] respectively.

**Figure 6:**
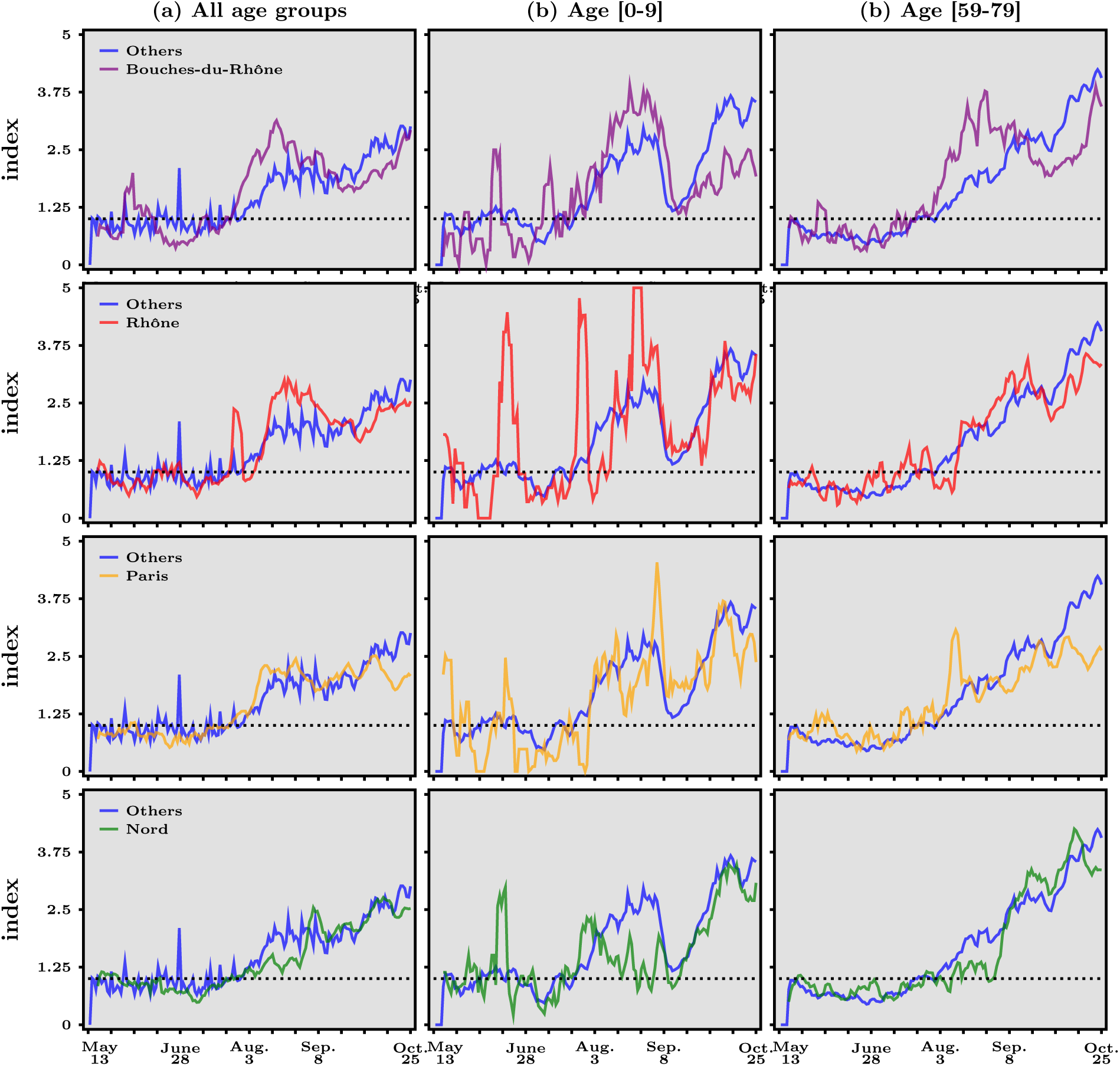
Acceleration index for the four largest French départements, May 13 - October 25, 2020. In each panel, the dashed horizontal line represents when acceleration index equals 1, the blue line depicts acceleration for all other départements while the colored lines correspond to the département specific to that panel (Bouches du Rhône, Rhône, Paris and Nord, from top line to bottom line). Data source: Agence Santé Publique France

In all panels, the blue line stands for the evolution of the acceleration index for all the other départements, while the second colored lines in each row represents the dynamics of acceleration for all age groups in panel (*a*) and the specific age group in panels (*b*) − (*c*). Looking first at panel (*a*), one sees that the dynamics in all four départements, resemble that of the others, with perhaps more volatility in the case of Bouches du Rhône and Rhône. Note, however, that the latter have experienced a period when the acceleration index decreased, roughly from mid-August to mid-September. This is not really the case for Paris and Nord.

In panel (*b*), we see considerable volatility, largely due to the fact that the absolute numbers of tests and positives are rather low for this age group at the level of the département, the acceleration index for the [0 − 9] age group hover around a plateau below one from mid-May to early July. What is striking, however, is that the acceleration index drops a lot before the end of August to levels close to unity, only to starts rising sharply again in the first half of September. As we asked before, why is there this rapid acceleration in this age group? Does it have to do with the dynamics of acceleration for the age group of grand-parents, depicted in panel (*c*). The rapid acceleration in this age-group is especially the case in Paris and Nord, where the acceleration index for the elderly rises continuously from the beginning of August, and may be less so for Bouches du Rhône and Rhône. In contrast, deceleration was pronounced both for kids and for grand-parents before the beginning of summer.

Although the above comments in relation to Figure 5 are merely descriptive, and deliberately so, they illustrate how our acceleration index can be used at any levels of granularity that is allowed by the available data and how it may also guide further analysis about transmission channels to complement contact tracing. An interesting avenue for research would obviously be to perform a panel data analysis, but this is beyond the scope of this paper.

## 3 An Acceleration-based Algorithm to Guide Test Strategies

Test strategies have many dimensions, including where to test and to allocate staff, locating groups at risk. We show in this section how to design an algorithm to allocate COVID-19 test resources. We focus, for the sake of illustration, on the spatial distribution of tests based on our acceleration index. Although testing is acknowledged as an efficient tool to curb epidemics^15^, little is known on the context of COVID-19 about *how* to geographically allocate a limited quantity of test.

Imagine now that public health authorities would like to decide how tests should be allocated to region *i* at date *T*. According to the decomposition stated in equation (1), a small increment of the per-period fraction of tests Δ*D*^*i*^ would be expected to lead to a proportional increase of positives 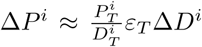 in region *i*. The logic of our criterion to allocate tests across regions has two steps. Firstly, we attribute a weight that equals 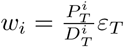 to each region *i*. Note that the weight *w*_*i*_ attributed to each region is the product of two terms: *(i)* the first term is the average rate of cumulated positives as a fraction of cumulated tests, at end date *T*, that is, 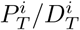; *(ii)* the second term is the acceleration index *ε*_*T*_, which measures whether the pandemic accelerates (i.e. *ε*_*T*_ > 1) or decelerates (i.e. *ε*_*T*_ < 1). This means that regions that are allocated more tests relative to others are those where either the overall positive rate is larger at end date, the pandemic accelerates more (or decelerates less), or both, relative to others. Second, we propose to allocate to region *i* a share of national test capacity that is given by:

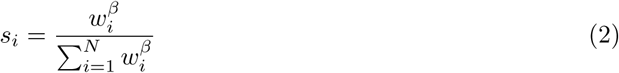

where *β* is a parameter which we assume takes positive real values: therefore, *s*_*i*_ goes up with *w*_*i*_ so that a region with a larger weight is allocated a larger share.

Our premise here is that any test strategy combines two aspects: *information* about the virus circulation and the *expected feedback* that testing implies, in terms of isolation and contact tracing most importantly. We therefore propose a parsimonious criterion to allocate tests across age groups and space, based on both our acceleration index and the positivity rate, and depending on the extent to which tests reduces the virus propagation. Because the latter feature cannot be measured with certainty, we argue that any allocation of tests should reflect the beliefs that public health authorities have about it and we therefore add a parameter to the analysis to take this key dimension explicitly into account. Obviously, we are assuming here a benevolent public health authorities, who act on objective grounds and are not trying to manipulate the indicator for political purposes.

Therefore, the allocation scheme that we propose builds upon the following logic. When *β* = 0, each and every region is allocated an *equal* fraction 1*/N* of the total amount of tests available at the national level. The other extreme configuration occurs when *β* tends to ∞ and it is easy to check that, in that extreme case, the region with the highest *w*_*i*_ is allocated *all* national tests.

One way to interpret our allocation rule for intermediate values of *β* is as follows. Parameter *β* measures the extent to which testing is believed to exert a *negative feedback effect* on the pandemic, for example because detecting positives entails isolating them and contact tracing, thereby reducing the number of contacts that produce additional contaminations. In the limit case when *β* = 0, there is no such feedback and a natural benchmark arises, which consists in allocating tests equally across regions. This corresponds to a situation where *information about the spatial distribution of the pandemic is maximized*, since each and every region has the same input in terms of testing. In this case, the objective is to gather information about where the pathogen is currently circulating. On the other hand, when *β* tends to ∞, all tests are allocated to the region with the highest *w*_*i*_ and this amounts to *maximizing the number of positives* so as to curb the pandemic, since in this case the negative feedback is believed to be the strongest.^16^ Middle cases happen when *β* is between zero and infinity, reflecting the belief held the public authorities deciding over the allocation of tests across regions about the strength of the feedback effect.^17^

We use Figure 7 to visually compare the allocation of tests, as computed from our algorithm based on the acceleration index, to the observed allocation and to the population distribution, as of October 25, 2020. In panel (*a*) and (*b*), we report the observed shares of tests and of total population in each département, respectively. Comparing the two upper panels reveals that the observed allocation of tests at that date reflects the geographical distribution of population to a large degree: in short, more tests have been done in more populated areas.

**Figure 7:**
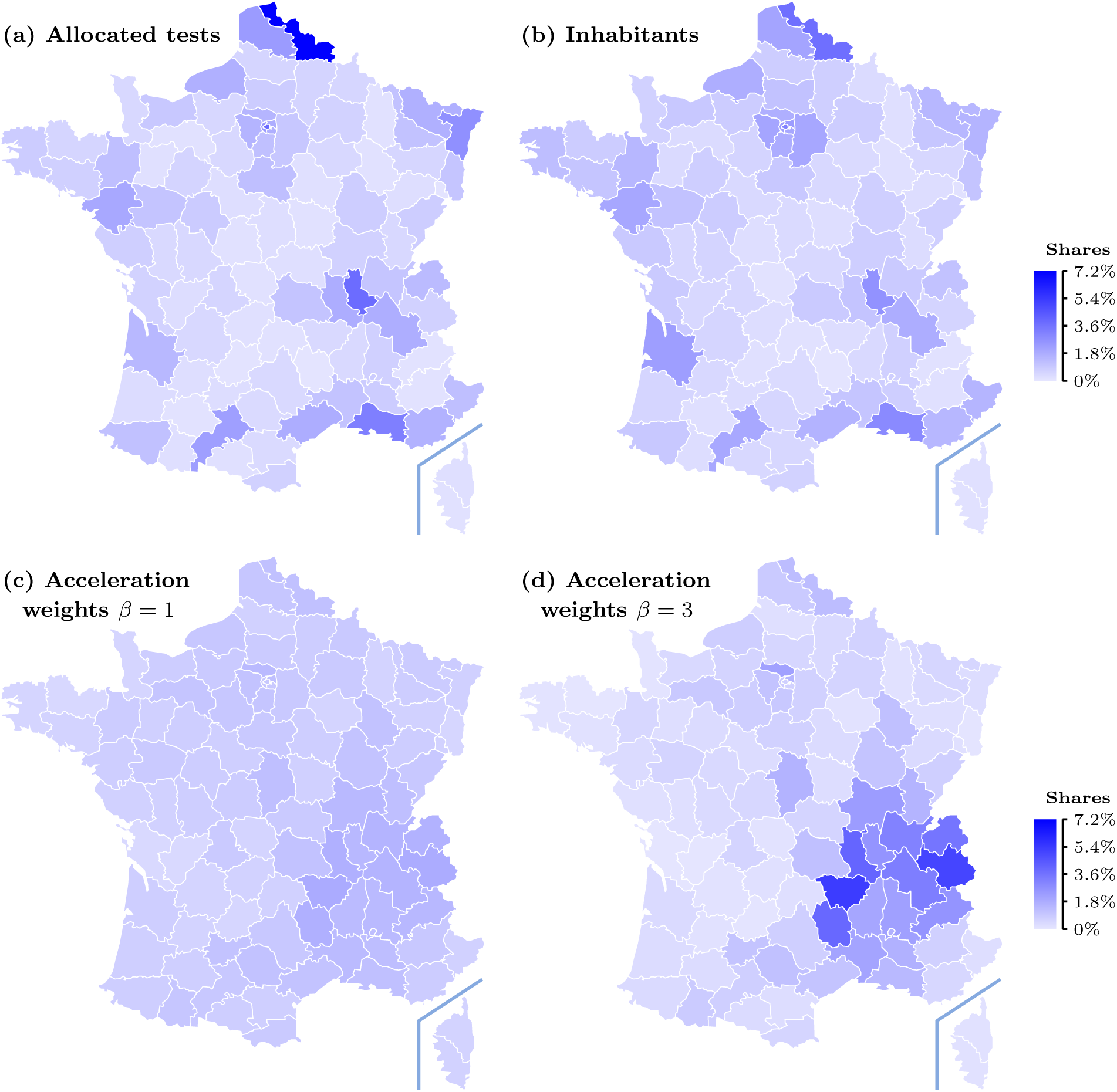
Allocation of tests and population in French départements, as of October 25, 2020 - actual in upper panels and allocation of tests derived from our acceleration index in lower panels. Data source: Agence Santé Publique France

Panels (*c*) and (*d*) in Figure 7 depict the allocation of tests over French départements, for different weights *β* put on the acceleration index. Comparing the upper and the lower panels makes clear that the spatial distribution of tests observed at October 25, 2020, has little connection to where the pandemic accelerates, which is somewhat paradoxical to the extent that detecting positives should help to curb the virus circulation. More precisely, while in panel (*a*) one sees that tests have been concentrated, especially in about roughly three sub-regions (Mediterranean South East, North, and Rhône), panels (*c*) − (*d*) shows that the allocation proposed by our algorithm based on the acceleration index is somewhat less concentrated or concentrated in different areas. This is particularly clear when *β* = 1. When *β* is higher, our algorithm allocates more tests where the acceleration index is larger than unity (and less tests where it is smaller than one) so that the allocation of tests becomes more concentrated but panel (*d*) shows how more tests should go to the south-east quarter of France. In contrast, panel (*a*) shows that this is not what has actually happened.

The difference between the actual distribution of tests over départements, which is very much population-driven, and our proposed allocation, which acceleration-centered can be established a it more formally by looking at the following measures. In Appendix D, we report in Tables 2-4 the numbers behind Figure 7 and we use those to compute two indicators that shed light on the differences between the actual and proposed spatial distributions of COVID-19 tests.

**Table 2:**
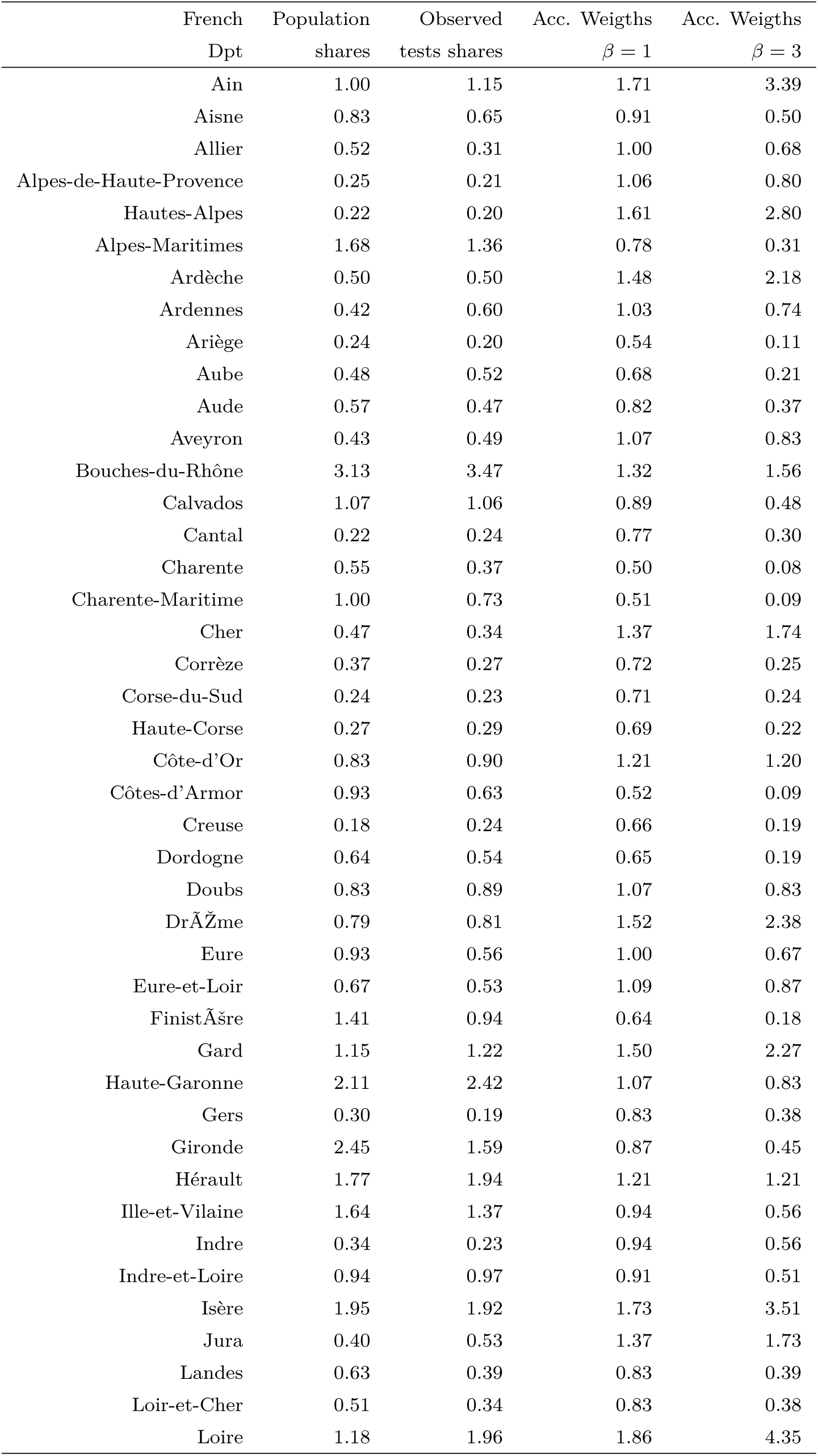
Allocation of tests in French départements - population vs acceleration, as of October 25, 2020. Data source: Agence Santé Publique France

**Table 3:** Allocation of tests in French départements - population vs acceleration, as of October 25, 2020 (Cted). Data source: Agence Santé Publique France

| French<br>Dpt | Population<br>shares | Observed<br>tests shares | Acc. Weights<br>$\beta = 1$ | Acc. Weights<br>$\beta = 3$ |
| --- | --- | --- | --- | --- |
| Haute-Loire | 0.35 | 0.50 | 2.04 | 5.71 |
| Loire-Atlantique | 2.16 | 2.13 | 0.86 | 0.42 |
| Loiret | 1.05 | 1.36 | 1.04 | 0.75 |
| Lot | 0.27 | 0.18 | 0.58 | 0.13 |
| Lot-et-Garonne | 0.51 | 0.33 | 0.70 | 0.23 |
| Lozère | 0.12 | 0.14 | 1.84 | 4.23 |
| Maine-et-Loire | 1.26 | 1.18 | 0.99 | 0.66 |
| Manche | 0.77 | 0.54 | 0.63 | 0.17 |
| Marne | 0.88 | 0.75 | 0.99 | 0.66 |
| Haute-Marne | 0.27 | 0.22 | 1.26 | 1.34 |
| Mayenne | 0.48 | 0.27 | 0.80 | 0.35 |
| Meurthe-et-Moselle | 1.13 | 1.27 | 0.89 | 0.47 |
| Meuse | 0.29 | 0.24 | 0.70 | 0.24 |
| Morbihan | 1.16 | 0.80 | 0.71 | 0.24 |
| Moselle | 1.61 | 1.84 | 0.96 | 0.61 |
| Nièvre | 0.32 | 0.22 | 0.81 | 0.36 |
| Nord | 4.03 | 7.70 | 1.29 | 1.44 |
| Oise | 1.28 | 0.95 | 0.96 | 0.60 |
| Orne | 0.44 | 0.24 | 1.16 | 1.04 |
| Pas-de-Calais | 2.27 | 2.57 | 1.14 | 1.00 |
| Puy-de-Dôme | 1.01 | 1.20 | 1.25 | 1.32 |
| Pyrénées-Atlantiques | 1.05 | 1.29 | 0.87 | 0.45 |
| Hautes-Pyrénées | 0.35 | 0.30 | 1.13 | 0.98 |
| Pyrénées-Orientales | 0.73 | 0.77 | 0.98 | 0.63 |
| Bas-Rhin | 1.74 | 2.92 | 0.81 | 0.35 |
| Haut-Rhin | 1.18 | 1.17 | 0.58 | 0.13 |
| Circonscription dép. du Rhône | 2.85 | 4.12 | 1.60 | 2.79 |
| Haute-Saône | 0.37 | 0.32 | 0.89 | 0.47 |
| Saône-et-Loire | 0.86 | 0.77 | 1.55 | 2.53 |
| Sarthe | 0.88 | 0.65 | 0.87 | 0.45 |
| Savoie | 0.67 | 0.75 | 2.00 | 5.39 |
| Haute-Savoie | 1.25 | 1.50 | 1.78 | 3.82 |
| Paris | 3.38 | 4.15 | 1.06 | 0.80 |
| Seine-Maritime | 1.94 | 1.86 | 1.09 | 0.87 |
| Seine-et-Marne | 2.17 | 1.09 | 1.04 | 0.75 |
| Yvelines | 2.23 | 1.71 | 1.22 | 1.22 |
| Deux-Sèvres | 0.58 | 0.50 | 0.77 | 0.30 |
| Somme | 0.89 | 0.62 | 0.76 | 0.29 |
| Tarn | 0.60 | 0.46 | 1.24 | 1.30 |
| Tarn-et-Garonne | 0.40 | 0.36 | 1.22 | 1.21 |
| Var | 1.64 | 1.78 | 0.94 | 0.56 |
| Vaucluse | 0.87 | 0.93 | 1.39 | 1.82 |
| Vendée | 1.04 | 0.82 | 0.75 | 0.28 |

**Table 4:** Allocation of tests in French départements - population vs acceleration, as of October 25, 2020 (Cted). Data source: Agence Santé Publique France

| French Dpt | Population shares | Observed tests shares | Acc. Weights $\beta = 1$ | Acc. Weights $\beta = 3$ |
| --- | --- | --- | --- | --- |
| Vienne | 0.68 | 0.54 | 0.88 | 0.46 |
| Haute-Vienne | 0.58 | 0.61 | 1.00 | 0.68 |
| Vosges | 0.57 | 0.59 | 0.77 | 0.31 |
| Yonne | 0.52 | 0.26 | 0.87 | 0.44 |
| Territoire de Belfort | 0.22 | 0.15 | 0.72 | 0.25 |
| Essonne | 2.01 | 1.34 | 1.17 | 1.07 |
| Hauts-de-Seine | 2.49 | 2.71 | 1.13 | 0.97 |
| Seine-Saint-Denis | 2.51 | 1.97 | 1.36 | 1.69 |
| Val-de-Marne | 2.15 | 1.90 | 1.04 | 0.77 |
| Val-d'Oise | 1.90 | 1.68 | 1.52 | 2.37 |

We first carry out a comparison between the different distributions using Jensen-Shannon (JS) divergence, which is based on Kullback-Leibler divergence and is normalized between 0 and 1, to measure similarities between the actual distribution of tests, population shares and our acceleration-based allocaton of tests. From Tables 2-4 in Appendix D, we find that JS divergence between the observed distribution of tests and the distribution of population equals 0.016, hence very small. This confirms our suggestion that relative population size has been an important determinant of the distribution of tests across French départements. In contrast, JS divergence between the observed distribution of tests and our proposed distribution based on acceleration is higher: it equals 0.115 when *β* = 1 and 0.194 when *β* = 3.^18^ Not surprisingly in view of the above discussion in relation to Figure 7, we conclude that observed and proposed distributions differ in a quantitative sense, but also that putting more weight on acceleration - that is, increasing *β* - further enlarges thus discrepancy.

Obviously, the allocation of tests and the pandemic’s dynamics are both endogenous and interactions between the two form a complex system. Our analysis is so far limited to tracking such dynamics and proposing an algorithm to allocated tests across space, according to the acceleration index. As such, it has little to say about the extent to which testing impacts negatively the virus circulation. For instance, it is reasonable to think the weights our algorithm put on acceleration should necessarily be the same in all départements. Perhaps isolation and contact tracing is more efficient in some regions that in others. In addition, additional constraints should arguably be introduced, such as the fact that the number of tests allocated to a département should not be larger than its population.^19^

Despite all these limitations, we still argue that our algorithm to allocate tests according to where the pandemic accelerates could be a valuable input to public health policies, not only in France of course but also in all countries facing COVID-19. Of particular importance, at this point, is how to reallocate the national testing resources and effort within département to specific age groups, and of course when the pandemic will start to decelerate, which one hopes is not too far ahead in time.

## 4 Conclusion

The purpose of this paper is to show that plotting the number of cases, of tests and even the positivity rate against time is far from the best way to measure the acceleration/deceleration of the virus. We propose instead a simple yet novel way to look at the data in real time. Our premise is that looking solely at the number of cases over time to measure acceleration of the pandemic is not accurate and hence problematic as a foundation for public health policies. This is because the amount of tests is far from constant per unit of time and this for a number of reasons. We thus argue that a much better understanding is gained by plotting the number of cases *against* the number of tests, that is, a simple and plain scatter-plot.

From such a scatter-plot we derive an acceleration index, which we propose, is a useful indicator to track the dynamics of pandemics like COVID-19 in real-time. Using French data on confirmed cases and tests for the period following the first lock-down - from May 13, 2020, onwards - our acceleration index shows that the ongoing pandemic resurgence can be dated to begin around July 7. Rather surprisingly, it helps to underline the fact that the pandemic acceleration has been stronger than national average for the [59 − 68] and especially the 69 and older age groups since early September, the latter being associated with the strongest acceleration index, as of October 25. In contrast, acceleration among the [19 − 28] age group is the lowest and is about half that of the [69 − 78]. We also propose an algorithm to allocate tests among French départements, based on both the acceleration of the pandemic and the feedback effect of testing. Our acceleration-based allocation differs significantly from the actual distribution over French territories, which is population-based.

We would like to stress that our approach has admittedly limitations, mostly due to the fact that it relies on actual data only and abstracts away from any theoretical model like those that have been stressed and used by some governments from early on. Yet, we do so deliberately and this for two reasons. First and foremost, COVID-19 has clearly put at the forefront the importance of admitting that the novelty of a new pathogen has implications for modelling. Any estimates obtained through, say, SIR or any other models, are subject to considerable and often unknown parameter uncertainty. This problem is particularly acute in the face of a novel pathogen, when public health authorities must act swiftly to prevent the virus from spreading in an uncontrolled way. Our analysis offers a real-time analysis that builds only on raw data and is, as such, subject to data measurement issues as any other approaches, but not to parameter uncertainty. Second, our real-time analysis is scale-free and can therefore be used at very granular levels, say cities or counties or age groups, where any model becomes rapidly either untractable or not transparent enough. In that sense, we think that our acceleration index carries important information, for instance compared to the ubiquitous reproduction numbers *R*_*0*_ and *R*_*e*_. In fact, the results in this paper could be used to go in the direction of advocating more diversity in the tools that should be used in an emergency crisis, that is, a type of “method averaging” (in the same sense that model-averaging is used) that seems to us necessary to take the best policy actions given the available information.

The index that we have exposed in this paper could also be used in the following weeks to monitor the pandemic and to detect positive signs of deceleration and to respond rapidly to new accelerations. The point is that our acceleration index might better track any reversal than looking solely at the average or daily positivity rate. When deceleration is under way, our index gives a very clear target for an all-clear: the acceleration rate has to be lower than one independently of where we look through our “acceleration lenses”, that is a spatial and group based analysis for example. It is also in this sense that our acceleration index can help assessing and modulating health policies to respond to those areas or groups where an acceleration takes place. Arguably, this would help avoiding generalized lock-downs, which are extremely costly, or other policies that affect the general society independently of whether they face an acceleration or not.

## Data Availability

Data used in the paper is public:
https://www.data.gouv.fr/fr/datasets/donnees-relatives-aux-resultats-des-tests-virologiques-covid-19/

## A Decomposing the Detection Effect of Tests

A country is divided into *N* regions, each indexed by *i* = 1, …, *N*. For each region, data is available about the number of tested and positive persons, up to end date *T*. Denote 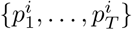 the historical times series of the new (per period) number of positive persons from date *t* = 1 to end date *t* = *T*. Similarly, 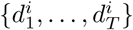 is the historical times series of new (per period) diagnosed/tested persons.

Denote 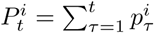 and 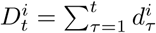 the cumulative numbers of positive and diagnosed persons up to date *t*. Finally, define 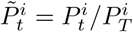 and 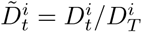 for *t* = 1, …, *T*, which are the fractions of, respectively, positive and diagnosed persons at date *t* relative to that at end date. In more technical term, dividing the historical times series by the most recent entry amounts in our setup with non-negative numbers to perform min-max normalization (see See Han, Kalber, and Pei [7], section 3.5.2). Our object of interest is the relationship between 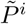 and 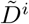 over time in the context of a pandemic, when testing is the only way to detect confirmed cases, which is depicted in the scatter-plot of Figure 2.

Suppose that the data 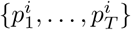 and 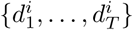 are used to estimate for each region a function *f* ^*i*^ such that 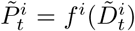 for *t* = 1, …, *T*. Because by definition 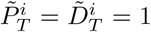 one has *f* ^*i*^(1) = 1. A first-order Taylor expansion at 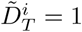 gives:

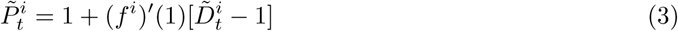

Equation (3) shows that the derivative (*f* ^*i*^)′(1) is essentially an elasticity, given the normalization of cumulative numbers by end date values: it measures how many positives follow an increase in the number of tests at end date, as a fraction of end date values. For example, values such that (*f* ^*i*^)′(1) > 1 mean that the pandemic is accelerating, since a given fraction of the total of tests performed up to date *T* is associated with a *larger* fraction of positives who are detected with those tests, in percentage of the cumulative number of positives at *T*. On the contrary, (*f* ^*i*^)′(1) < 1 implies that the pandemic is decelerating. This elasticity is what we have labeled the acceleration index and labeled *ε*_*T*_ in Section 2.1.

All of the above implies that, given the estimate of the first-order derivative, *ε*_*T*_ ≡ (*f* ^*i*^)′(1), equation (3) can be rewritten in terms of the numbers of positive and tested persons, that is:

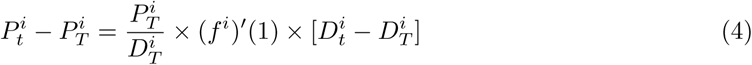

In other words, equation (4) can be used to decompose the effect of tests on positives *in levels*, that is, how many additional positives are detected given additional tests, between *T* and *T* + *dt*:

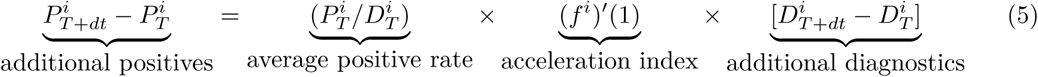

Equation 5 is identical to equation 1 in Section 2.1, where *ε*_T_ is estimated as the ratio of variations of cases and tests between *T* and *T* − 1. Similarly, the above decomposition in levels holds for any date *t* < *T*, as follows:

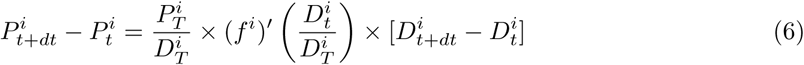

From equation (6), the effect of tests on positives in percentage terms from the perspective of date *t* is therefore written as:

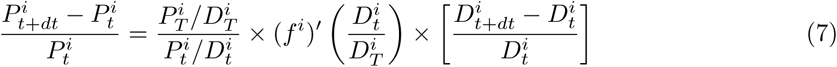

The elasticity of the number of positives with respect to the number of tests is now, because it is evaluated at date *t* as opposed to end date *T*, the product of the derivative at the relevant point, times the ratio of average positive rates – that of date *T* over that of date *t*.

## B Decomposition and Acceleration index: Exponential Case

This section explores what the decomposition stated in Section A reveals when time is assumed to be continuous and when the number of cases grows exponentially over time, as usually assumed in epidemiological models, of SIR type and related for example. Although typically absent in the latter strand of literature, we have to introduce tests and we assume that they also grow exponentially. More formally, using the notation in the previous section, suppose that the number of cases per unit of time is denoted by *p*(*t*) = *αe*^*βt*^ while the number of tests per unit of time is *d*(*t*) = *γe*^*νt*^, where the growth rates *β* and *ν* are assumed to be positive for the sake of illustration. Cumulated cases and tests are then noted 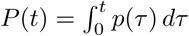 and 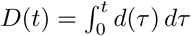, respectively. It is easy to derive, by straight integration, the expressions:

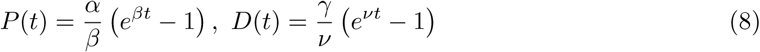

It follows that our acceleration index is given, as function of time, by:

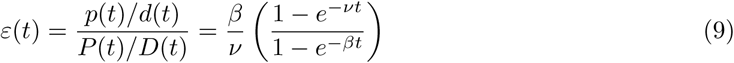

From equation (9), then, one infers that two cases occur. When *β* = *ν*, that is, when both cases and tests grow at the exact same rate, then our acceleration index equals 1 at all dates. When the two growth rates differ, however, *ε*(*t*) converges, when *t* goes to infinity, to the ratio of growth rates *β/ν*, independently of the scale parameters *α* and *γ*. As an illustrative example, suppose that *β* > *ν*, so that positives grow faster than tests. Then the pattern of our acceleration index *ε*(*t*) over time will have two regimes: it first grows almost linearly and eventually reaches the upper bound *β/ν* > 1. Obviously, in that case both the daily positivity rate *p*(*t*)*/d*(*t*) and the average positivity *P* (*t*)*/D*(*t*) grow over time, and the latter quantity exceeds the former all the time so that acceleration prevails. This closely resembles the pattern following early August to early October in Figure 3, as underlined in the main text.

## C Statistics for Age Groups

In Table 1 we report a few statistics for all age groups, as of October 25, 2020. In the second and third columns we report the numbers of cumulated cases and tests, respectively. The fourth column depicts that average positivity rate, defined as the ratio of cumulated cases and cumulated tests, while the actual test shares appear in the fifth column. Finally, the last column shows the share of cases by age group, which is defined as the ratio of cumulated cases.

## D Test Allocation across Départements: Actual vs Acceleration-based

## Footnotes

1 The Spanish newspaper “El Pais” has investigated the data-problem in Spain for example. See https://english.elpais.com/society/2020-06-24/the-problems-spains-outdated-data-methods-have-caused-during-a-21st-century-pandemic.html?rel=mas.

2 See Gurdasani and Ziauddeen [6] for an example.

3 We focus on the post-lockdown period in France - starting May 13, 2020 - since too few tests had been performed before that date. As a consequence, the estimated numbers of positives are not reliable before the end of the general lock-down. This is probably the case for many other countries, but not for a few others like South Korea.

4 All the data used in this paper is publicly available from the web page “Données relatives aux résultats des tests virologiques COVID-19 SI-DEP” https://www.data.gouv.fr/fr/datasets/donnees-relatives-aux-resultats-des-tests-virologiques-covid-19/.

5 Santé Publique France decided at some point to report also the positivity rate, that is, the ratio of positive cases to tests at weekly frequency. However, as we will argue, even this indicator does not convey enough information and possible early warning signals that public health authorities could rely on.

6 An important reason behind the observed non-constancy of tests over time in most countries is undoubtedly the fact that the pandemic caught most of the world by surprise in the first quarter of 2020. As a consequence, the widespread goal to progressively increase the capacity to perform tests on a large scale has led many countries to increase the number of tests since March.

7 See https://www.who.int/dg/speeches/detail/who-director-general-s-opening-remarks-at-the-media-briefing-oncovid-19-16—march-2020 last accessed April 14th 2020.

8 Guidelines from the ECDC explicitly state that the presence of SARS-Cov2 should be assessed via tests and that only cases confirmed that way should be counted in the official statistics. See https://www.ecdc.europa.eu/sites/default/files/documents/TestingStrategyObjective-Sept-2020.pdf

9 See Han, Kalber, and Pei [7], section 3.5.2. Broadly speaking, min-max normalization preserves the relationships, such as quantiles for example, among the raw data before normalization.

10 More generally, the (local) elasticity of a given function is the ratio between its derivative and its average value, taken at a particular point. See Arrow et al. [1]. The empirical counterpart that we use in this paper is conceptually similar.

11 Mathematically speaking, the normalization and updating of the data means that data points are subject to a rotational homothety over time. For instance, while the end point at *T* has coordinates (1, 1), it is subject at date *T* + 1 to a mapping which is a composition of a contraction, due to the new values of cases and tests at end point, and of a rotation, due to the new value of the ratio between the latter. This is why the colored points move over time in Figure 2.

12 Our acceleration index has, to our knowledge, not been studied before, but many health agencies around the world report the positivity rate over time. In particular, Santé Publique France started, during the second quarter of 2020, to report the *weekly* positivity rate on top of the number of cases. The first time a rise of such an indicator is noted is July 23, in the “Point épidémiologique hebdomadaire du 23 juillet 2020”, whereas our acceleration index starts rising on July 7 as shown above. This two-week gap may suggest that using daily data rather than weekly data is more appropriate.

13 We conjecture that such a global measure of convexity relates to *δ*-convexity as defined by Hyers and Ulam, 1952.

14 https://www.ecdc.europa.eu/en/covid-19/situation-updates/weekly-maps-coordinated-restriction-free-movement

15 The importance of testing is most clearly spelled out in an official document that has been produced by the South Korean government for the international community. See “Flattening the curve on COVID-19 - How Korea responded to a pandemic using ICT”, dated April 15, 2020.

16 In reinforcement learning terminology, that would lead to stop exploring others aera and only to exploit the region with highest *w*_*i*_ when the algorithm starts much like a *greedy* algorithm would do - see Sutton and Barto [10]. This is however not a desirable property in the case of a pandemic and acceleration weights should always lead to *some* exploration to gather information about the virus circulation in other areas, such as in *ϵ*-greedy algorithms.

17 Note that in theory, *β* could be negative. This would the case, for instance, if positives would have even more contacts with susceptible persons, generating even more contaminations, compared to a situation with no test. In this case, the share of tests *a*_*i*_ would go down when *w*_*i*_ goes up. Although such a situation is hypothetically possible – think about bio-terrorism – we rule it out for the sake of realism.

18 Of course, JS divergence between the distribution of population and our proposed distribution features close values: it equals 0.092 when *β* = 1 and 0.193 when *β* = 3.

19 While such constraint is unlikely to bind at the département level, it could at more granular levels at which our algorithm could operate as well. For example, some little populated cities could be allocated too much tests in view of their populations. In that case, our algorithm could easily be amended to account for such constraints, by capping test and reallocating the surplus of test according to the next cities where acceleration is the highest.

